# Dopaminergic-related Anatomical Pattern of Dorsal Striatum in Schizophrenia

**DOI:** 10.1101/2023.11.30.23298956

**Authors:** Chao Xie, Shitong Xiang, Yueyuan Zheng, Chun Shen, Xuerui Peng, Yuzhu Li, Wei Cheng, Xiao Chang, Jingliang Cheng, Long-Biao Cui, Chu-Chung Huang, Nanyu Kuang, Chunbo Li, Ching-Po Lin, Cheng Luo, Yingying Tang, Jijun Wang, Xinran Wu, Dezhong Yao, Jie Zhang, Tianhong Zhang, Andreas Heinz, Trevor W. Robbins, Oliver D. Howes, Gunter Schumann, Tianye Jia, Jianfeng Feng

## Abstract

Striatal dopaminergic overactivity was hypothesized as the core pathophysiology of schizophrenia. However, morphological alterations of striatum in schizophrenia remains exclusive, largely because brain regional heterogeneity limited traditional group-mean based approach. Leveraging third-party brain maps of neurotransmitter and cognition behaviours, we developed a pattern-based representation feature score (ReFS) to investigate structural spatial pattern variation in schizophrenia. Structural ReFS of subcortical regions, particularly the striatum, were linked to schizophrenia diagnosis, symptom severity, and genetic susceptibility. Dopaminergic-ReFS of striatum was increased in schizophrenia patients and reliably reproduced across 13 datasets. The pattern-based ReFS effectively captured the shared genetic pathways underlying both schizophrenia and striatum. The results provide convergent, multimodal suggest the central role of striatal spatial patterns in schizophrenia psychopathologies and and open new avenues to develop individualized treatments for psychotic disorders.

## Introduction

Schizophrenia and related psychotic disorders are major contributors to worldwide disease burden, which is partly due to the lack of efficacy of antipsychotic drugs for many patients, as well as significant side effects caused by the medication (*1*). Both shortcomings are likely related to an insufficient understanding of the neurobiological mechanisms underlying the disorder. Thus there is a need to improve understanding of the pathophysiology of schizophrenia to help develop better treatments (*2*). Several lines of in vivo imaging evidence suggest that increased striatal dopamine synthesis and release capacity underlie psychotic symptoms (*3*),,including across disorders (*4*). While these studies identify the striatum as a key brain region in the etiology of psychotic disorders, a comprehensive understanding of striatal abnormalities beyond increased dopamine release remains elusive (*5*). In particular, the evidence for striatal volume alterations in schizophrenia is inconsistent (*6, 7*).

One reason for this inconsistency could be because group mean-based gray matter volume of brain regions has commonly been used to examine the structural differences in schizophrenia, based on the implicit assumption that there is homogeneity in the measure between groups within each region of interest. However, mounting evidence suggests there is greater heterogeneity among structural measures in schizophrenia patients compared to controls (*8*). Moreover, other evidence shows that levels and activity of neurotransmitter systems varied spatial distribution within brain regions, and that people with schizophrenia show differences in this which are missed by regional comparisons (*9, 10*). These findings indicate that the region-based approaches to analysis brain data miss intricate, high-dimensional spatial information within the brain that are relevant to understanding mental disorders such as schizophrenia(*11, 12*).

To address this, we aimed to develop and test a reference-based method that analysed the pattern of variation in brain structural data (**Fig S1**). We decomposed the complex spatial information of brain structure in MRI scans by transforming region-level spatial pattern of grey matter volume into a multidimensional representational space that was constructed using a specific third-party functional reference map (**Fig. 1a**). Specifically, for each brain region, we normalized individual-level grey matter volume (GMV) and group-level functional reference maps, followed by calculating the paired-wise spatial similarity for each individual. This process generates representational feature scores (ReFS) of latent brain functions for each brain region. We then tested and demonstrated that the spatial reference was highly reliable across datasets, and the structural ReFS has excellent reproducibility across scanner types, sequences, and time points, as well as a high sensitivity to individual differences (**Table S1, Fig. 1b and Fig S3-6**). Next, we discovered that structural ReFS of subcortical regions, particularly the striatum, were linked to schizophrenia diagnosis, symptom severity, and genetic susceptibility (**Fig S2**). Finally, we revealed that it was the reference-based ReFS, rather than mean-based volume based on region of interest analysis, that effectively captured the shared genetic pathways underlying both schizophrenia and brain structure.

**Fig 1.**
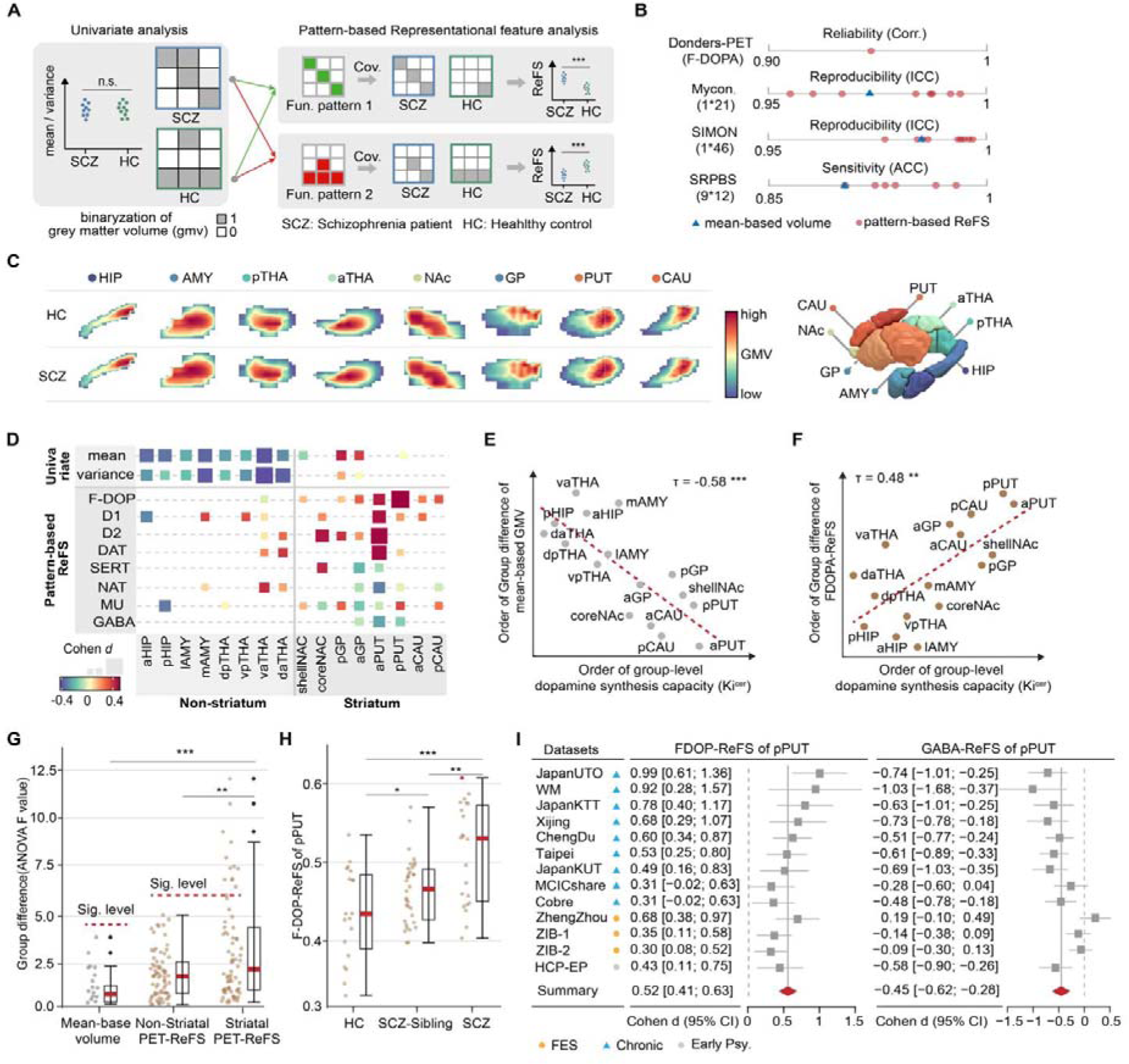
The structural pattern-based representation feature analysis of striatum in schizophrenia. **a.** Concept of pattern-based representation feature analysis. In regions with high spatial heterogeneity, univariate analysis with mean/variance-based grey matter volume (GMV) failed to find significant group difference between schizophrenia patients (SCZ) and healthy controls (HC). Spatial organization of functional references provide an priori knowledge to decompose the spatial pattern of GMV into different latent representation dimensions, which was termed as representational feature score (ReFS). The pattern-based ReFS help to detect the subtle spatial distribution alternations of brain structure. **b**. The reliability, reproducebility and sensitivity analysis results of the ReFS and mean-based volume. For each region, we used nine different neurotransmitter maps as pattern reference to estimate the pattern-based ReFS, such as FDOP-ReFS, D1-ReFS etc. With the F-DOPA as an exmaple, the reliability of brain ReFS was estimated as the spatial correlation between the F-DOPA maps from two public available third-party datasets (Donders dataset and JuSpace). The reproducebility of brain ReFS and mean-based volume was estimated as the intraclass correlation coefficient (ICC) with two repeated scanning datasets (Myconnectome dataset, 1 participant was scanned 21 times; SIMON dataset, 1 participant was scanned 46 times). The sensitivity of brain ReFS and mean-based volume was estimated as the accuracy in the brain fingerprint analysis with a participant travelling dataset (SRPBS, 9 participants were scanned at 12 different sites). **c**. For each subcortical region, there are complex nuanced group difference of spatial pattern between schizophrenia patients and healthy controls. d. The group difference between schizophrenia and healthy control groups with univariate-based measurements of mean and variance and patter-based ReFS, which was estimated with nine group-level neurotransmitter maps. **e.** In subcortex, the group difference of schizophrenia with mean-based volume was negatively associated with the group-level dopamine synthesis capacity (F-DOPA). **f**. In subcortex, the group difference of schizophrenia with F-DOPA-ReFS positively correlated with group-level dopamine synthesis capacity (F-DOPA). **g.** After multiple corrections with Benjamin–Hochberg procedure, subcortical region with mean-based volume did not show significant group difference between schizophrenia patients, unaffected siblings, and healthy controls; in contrast, the group difference with multiple PET-ReFS of striatum reached significant level, including F-DOPA-ReFS of pPUI, D2-ReFS of pPUT, MU-ReFS of pGP. In subcortex, the group difference with PET-ReFS exhibit significant higher level than with mean-based volume. The significance level was set as the corresponding multiple correction in 16 regional mean-based volumes and 144 (16 regions * 9 neurotransmitters) pattern-based subcortical PET-ReFS. The upper and lower bars represent the Q3 + 1.5 × IQR and Q1 − 1.5 × IQR, respectively. The upper and lower edges of a box represent the Q3 and Q1, and the central line represents the median. Outliers are illustrated as bold dots. h. F-DOPA-ReFS of pPUT demonstrates a significant genetic risk gradient from healthy controls, through unaffected siblings, to schizophrenia patients. **i.** Forest plot showing the effect sizes for the F-DOPA-ReFS and GABA-ReFS of pPUT between schizophrenia patients and healthy controls. Meta-analysis showed there was significantly higher F-DOPA-ReFS for pPUT (cohen’s d=0.52 (95%CI = [0.41, 0.63]); P < 0.0001), whilst it was significantly lower for GABA-ReFS for o=pPUT in schizophrenia relative to controls (cohen’s d=--0.45 (95% CI= [-0.62 −0.28]); P < 0.0001). Abbreviation: *SCZ*, schizophrenia patient; *HC*, healthy control; *ICC*, intraclass correlation coefficient; *F-DOP*, Fluorodopa; *FES,* First episode schizophrenia patient; *Chronic*: Chronic schizophrenia patient; Early Psy. Early psychosis patient. Sig. level, Significant level after multiple correction; * P < 0.05, ** P < 0.01, *** P < 0.001. Q1, first quartile; Q3, third quartile; IQR, interquartile range; SIMON, The Single Individual volunteer for Multiple Observations across Networks dataset; SRPBS, Japanese Strategic Research Program for the Promotion of Brain Science. aHIP, anterior hippocampus; pHIP, posterior hippocampus; lAMY, lateral amygdala; mAMY, medial amygdala; vpTHA, ventral posterior thalamus; vaTHA, ventral anterior thalams; dpTHA, dorsal posterior thalamus; daTHA, dorsal anterior thalamus; coreNAC, core nucleus accumbens; shellNAC, shell nucleus accumbens; aGP; pGP; aPUT; pPUT; aCAU; pCAU; FDOP; GABA; D1, D2, SERT, MU, NAT; DAT.

## Results

### Altered representation feature scores (ReFS) of striatum structure in schizophrenia

As neurotransmitter receptors are central to the propagation of signals in the human brain, we first estimated brain structural representational feature scores based on neurotransmitter maps derived from positron emission tomography (termed as PET-ReFS, **Fig S7**). We found that in a large-scale multisite schizophrenia dataset (1,229 SCZ participants, 1,237 HC participants, **Table S2**), the Schizophrenia group showed significantly higher dopamine-ReFS of striatal regions, including posterior putamen (pPUT), anterior putamen (aPUT) and core Nucleus Accumbens (coreNAC) (**Fig. 1c, Table S3**). Besides, we also found that schizophrenia patients showed lower gamma-aminobutyric acid (GABAa)-ReFS in aPUT and pPUT (**Fig. 1c, Table S3**). After statistically accounting for the influence of one another, the results of GABAa-ReFS and dopamine-ReFS remained significant (**Table S3**), suggesting that the two neurotransmitters have unique contributions to the neurobiological mechanisms of schizophrenia.

Whereas in the striatum we found differences in ReFS, but not in mean-based GMV, significant group differences in regional mean and variance of grey matter volume (GMV) were apparent in non-striatal regions (**Fig. 1c and Table S3**), especially the hippocampus (HIP) and thalamus (THA), which is consistent with previous mean-based GMV studies (*6, 13*). We further revealed that subcortical regions with higher dopamine synthesis capacity (F-DOPA) demonstrated less differentiated mean-based GMV (τ = −0.58, *P* < 0.001, **Fig. 1d**) but higher F-FOPA-ReFS abnormalities (τ = 0.48, *P* < 0.001, **Fig. 1e**) between schizophrenic patients and controls. The results suggest that the reference-based spatial pattern was more closer to schizophrenia etiology.

Moreover, with a dataset consisting of healthy controls, schizophrenia patients, and unaffected siblings, we observed that the PET-ReFS revealed greater group differences than for mean-based GMV, especially in the striatum (*t* = 2.37, *P* = 0.019, **Fig. 1f**) and the mean-based GMV of subcortical regions revealed no significant group differences (**Fig. 1f, Table S4**). For example, F-DOPA-ReFS in pPUT showed an apparent genetic susceptibility gradient for schizophrenia (*F*(_2,72_) = 10.64, *P* = 8.92E-05) (**Fig. 1g** and **Table S4**), which is consistent with previous finding that elevated dopamine synthesis capacity (F-DOPA update) in the dorsal striatum of subjects at ultra-high risk for psychosis (*14*). These results suggest that the pattern-based representational analysis is sensitive in detecting the morphological disruptions associated with dopaminergic alterations in schizophrenia, and structural ReFS serve as an intermediate structural phenotype linking brain structure and genetic mechanisms underlying schizophrenia.

Furthermore, the PET-ReFS showed consistent group differences across all 13 datasets (**Table S5**), as shown in Figure 1 (pPUT: Cohen’s *d 0.30∼ 0.99*; Overall: *t* = 10.62, Cohen’s *d* = 0.49, *P* = 8.17E-26; **Fig1. h**). Subsequent sensitivity analyses showed that the effect of F-DOPA-ReFS in pPUT was robust to clinical subgroups (first episode and chronic, **Fig1. h**), disease duration (long and short duration groups), sex (male and female groups), and age (young and old groups) (**Table S6**), while the effect of GABAa-ReFS in pPUT was observed in the chronic patients, but not in the first-episodic subgroup (**Fig 1. h** and **Table S6**).

In summary, these findings indicated that reference-based spatial pattern analysis exhibits high consistency and sensitivity, and it outperforms traditional univariate mean-based analysis in detecting fine-grained structural abnormalities within dopaminergic brain regions.

### Structural abnormality of dorsal striatal ReFS with latent cognitive factor in schizophrenia

Databases of brain activation maps provides a quantitative approach to infer plausible cognitive processes related to a spatial pattern, which might provide greater insight into the nature of the mental processes that are disrupted in the course of schizophrenia (*15*). We utilized meta-analytical decoding references from the *Neurosynth* dataset to transform individual-level spatial pattern of each subcortical region into a high-dimensional latent cognitive factor (termed as Cog-ReFS, **Fig S8**) (*16, 17*). Again, after controlling for multiple comparisons with the Benjamini–Hochberg procedure (16 region*539 cognitive factors), we found that the abnormalities of Cog-ReFS in schizophrenia patients were mainly located in dorsal striatal regions (63.4%, 1009/1591 significant Cog-ReFS), especially the posterior globus pallidus (pGP, 247 Cog-ReFS) and pPUT (274 Cog-ReFS) (**Fig. 2 a** and **Table S7**).

**Fig 2.**
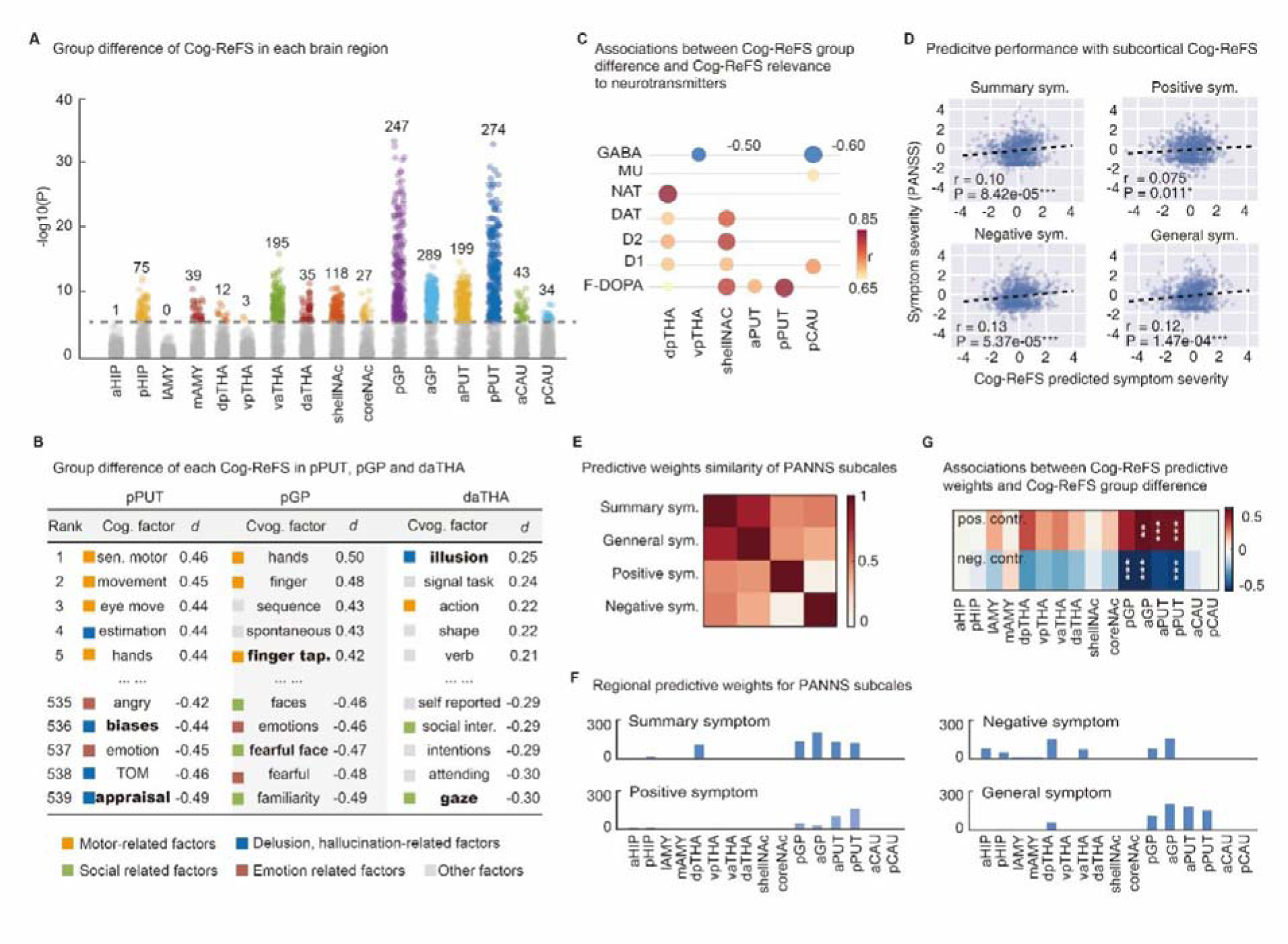
Striatal ReFS with multidimensional latent cognitive factors in schizophrenia patients. **a.** Manhattan plot illustrates significant group differences between schizophrenia and healthy controls in Cog-ReFS of subcortical regions, particularly in pGP and pPUT. **b**. The group difference of Cog-ReFS in pPUT, pGP and daTHA. **c**. In these regions, group differences of Cog-ReFS were notably modulated by neurotransmitters. **d**. High dimensional Cog-ReFS in subcortical regions could significantly predict the schizophrenic symptom subscales. **e**. The similarity between predictive weights for different schizophrenia symptom subscales. **f**. The regional contribution of subcortical Cog-ReFS in the predictive models of schizophrenia symptoms. **g.** The similarity between the summary symptom’s predictive contributors and case/control difference for all the Cog-ReFS in each subcortical region.

For instance, we observed significantly increased Cog-ReFS in pPUT with sensory-motor factors, such as “*eye movement*” and “*hand*”, as well as social factors and decisions, such as “*theory of mind*” and “*appraisal*” in schizophrenia patients (**Fig. 2b)**. Such an observation is in line with characteristic deficits or deteriorations in both motor and social factors of schizophrenia (*18*). Similarly, schizophrenia patients also showed a greater Cog-ReFS of motor-related factors (such as “*finger tapping*”) but a lower Cog-ReFS for emotional and social functions such as “*familiarity*” and “*fearful face*” in pGP (**Fig. 2b)**, where the “*finger tapping*” test is a commonly adopted measure of motor deficits in schizophrenia (*19*). In addition, for Cog-ReFS in the dorsal anterior thalamus (daTHA), the “*illusion*” term was the most prominent finding (**Fig. 2b**). Notably, delusions-related functions (”*biases*”, “*appraisal*”) are the hallmark positive symptoms of schizophrenia (*20, 21*).

Further, by investigating the correlations between the Cog-ReFS case/control differences and the third-party pattern similarity of cognitive factor maps and neurotransmitter maps (**Supplementary method and Fig S9**), we confirmed that the contribution of Cog-ReFS for schizophrenia could be partly explained by the neurotransmitters (**Fig. 2c, Table S8**). The above results altogether hence suggested that structural representational pattern could integrate cognitive factor and molecular neurotransmitter to delineate the neuropsychopathological mechanism of schizophrenia convergently.

We then investigated whether the high-dimensional Cog-ReFS in subcortical brain regions could predict the clinical characterisation of schizophrenia, as measured by the Positive and Negative Syndrome Scale (PANSS). Leveraging a modified linear predictive model (**Supplementary Methods and Fig S10**), we found that subcortical Cog-ReFS could significantly predict the summary symptom score and all three symptom subscales (Summary score: *r* = 0.10, *P* = 8.42E-05; Positive symptoms: *r* = 0.075, *P* = 0.011; Negative symptoms: *r* = 0.13, *P* = 5.37E-05; General symptoms: *r* = 0.12, *P* = 1.47E-04, **Fig. 2d**). We also found that the predictive weight of positive and negative subscale show significantly lower similarity (**Fig. 2e, Table S9**), suggesting that the latent cognitive factors mapping could effectively characterise the different neurobiological configurations of positive and negative schizophrenia symptoms. In terms of the regional predictive weights, the dorsal striatal regions again show predominant contributions, especially the GP and PUT (**Fig. 2f, Table S9**). At last, we show that only in aGP and pPUT, the positive and negative predictors of schizophrenia symptoms were significantly associated with the Cog-ReFS case/control differences (Table S7-c, **Fig. 2g and Fig S11, Table S9**). These results might indicate that the psychopathology of schizophrenia and the clinical symptoms of schizophrenia patients could share similar cognitive abnormality spectra centred at the dorsal striatum.

### Genetic links between structural ReFS and schizophrenia

Despite high heritability of both schizophrenia and striatal mean-based volumes, they were reported not to share similar genetic architectures(*22*). Given the above higher clinical relevance of reference-based spatial pattern for SCZ than mean-based volume, we last investigated whether structural ReFS with higher information specificity might help to bridge the gap between structural brain genetics and risk for schizophnreia.

Utilised imaging-genetic data of 34,497 participants of British ancestry from UK Biobank, we found the mean heritability (*h*^2^) of brain Cog-ReFS (8624, 16 regions* 539 cognitive factors) across the 16 subcortical regions as 30.1% (7.7%∼61.9%, all P<0.001, **Table S10 and Fig S12**). While the above heritability levels of Cog-ReFS are comparable with previous brain structural findings (*23, 24*), they are largely unchanged after regressing the corresponding brain volume (mean *h*^2^ = 28.6%, Difference heritability = 1.5%; individual heritability changes vary from −1.1% for vaTHA to 4.39% for aGP) (**Table S11 and Fig S12**). Hence, the subcortical Cog-ReFS may help to capture the remaining genetic variance of the subcortical structure that was largely missed by using volumetric measurements alone.

We then used a high-definition likelihood method to estimate the genetic correlation between schizophrenia and subcortical Cog-ReFS (*22, 25*) (**Fig. 3a left**). After controlling for multiple comparisons, the Cog-ReFS of all 16 subcortical regions had significant genetic correlations with schizophrenia (|Rg|: 0.022∼ 0.153, 2676 out of 8624 ReFS, Fig. 3b, **Fig. 3a right** and **Table S12**). Remarkably, using the Cog-ReFS of pPUT, we identified 116 risk Cog-terms (Rg 0.037∼0.089, mainly for sensorimotor and attentional factors) and 122 protective Cog-terms (Rg - 0.118∼-0.033, mainly for social and language factors) for schizophrenia (**Fig. 3b** and **Table S13**).

**Fig 3.**
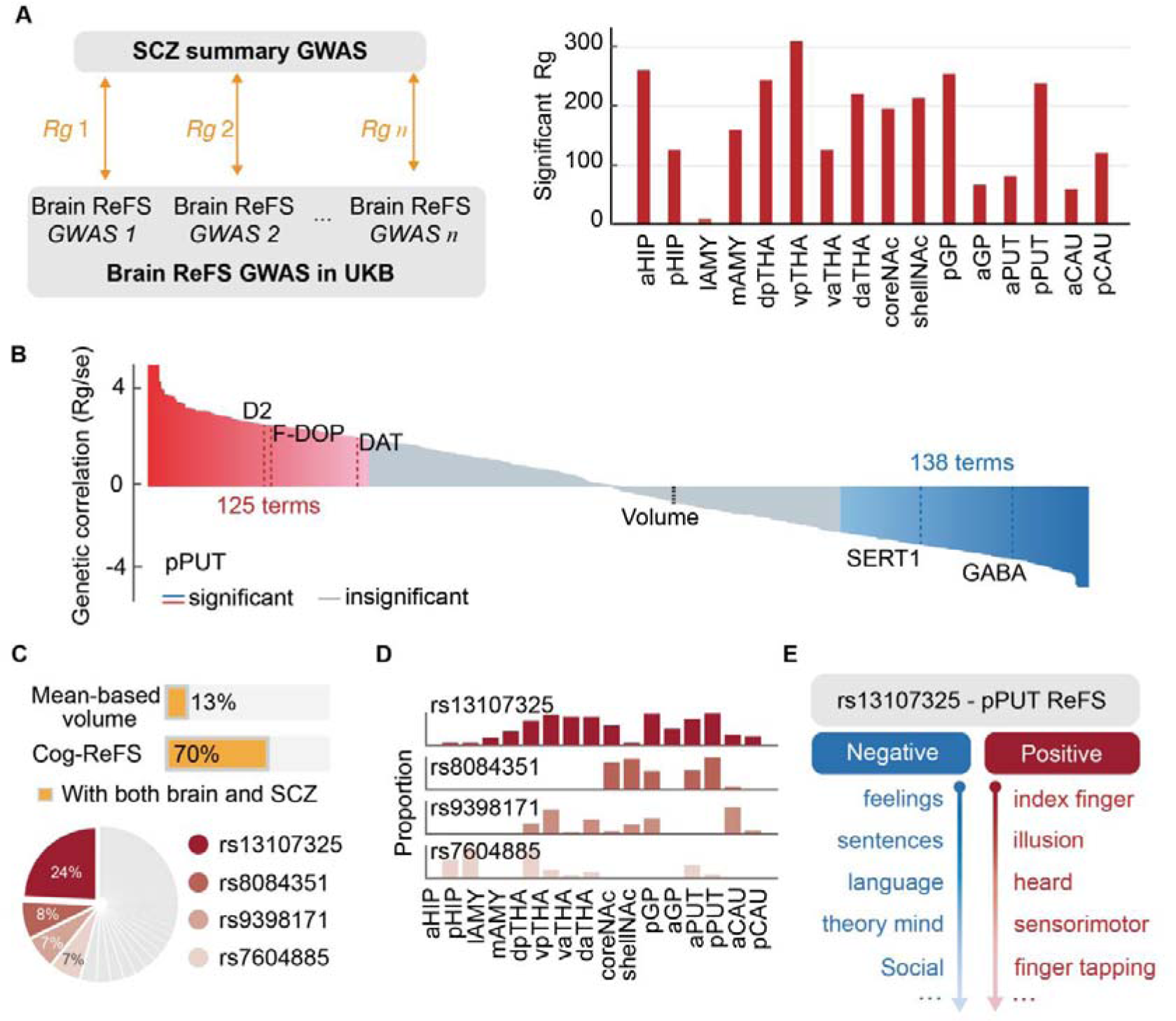
The shared genetic basis between brain Cog-ReFS and schizophrenia. **a.** The genetic correlations were calculated between brain ReFS (i.e. PET-ReFS and Cog-ReFS) of subcortical regions (GWAS using UKB) and the summary statistics from a recent schizophrenia GWAS (Left). The counts of significant findings are shown on the Right. **b**. In pPUT, ReFS with 125 latent cognitive factors and three neurotransmitters (D2, F-DOPA, DAT) had positive significant genetic correlations with schizophrenia, and 138 latent cognitive factors and two neurotransmitters (SERT1 and GABA) had negative genetic correlations. The volume of pPUT was not significantly associated with schizophrenia. **c**. Among the 279 single nucleotide polymorphisms (SNPs) from a recent schizophrenia GWAS, 70% of SNPs (172) have significant correlations with the subcortical ReFS after false discovery rate (FDR) correction and only 13% SNPs with subcortical mean-based volume. Four SNPs, i.e. rs13107325, rs8084351, rs9398171, and rs7604885, predominated the SNP-brain ReFS associations (46%). **d**. The SNP-brain ReFS associations of the four most representative SNPs showed different regional distributions. **e**. SNP rs13107325 showed negative associations with the Cog-ReFS in pPUT for social emotion and language terms and positive associations for motor and illusion terms. Rg, the genetic correlation; ReFS, representational feature score; GWAS, genome-wide association study; UKB, UK biobank

We also identified significant genetic correlations between the PET-ReFS of pPUT with schizophrenia, i.e. positive for the ReFS of D2 (dopamine D2), F-DOPA and DAT (dopamine transporter) and negative for the ReFS of SERT (serotonin transporter) and GABAa (**Fig. 3b** and **Table S14**). In contrast, the genetic correlation between volumetric pPUT and SCZ was not significant (Rg = −0.01, P = 0.46), consistent with previous studies (*26*) (**Fig. 3b** and **Table S15**). Further, also in pPUT, the magnitude of genetic correlations between Cog-ReFS and schizophrenia was significantly associated with the Cog-ReFS difference between schizophrenia patients and controls (*r* = 0.84, *P _perm_* = 0.01, Fig S14), hence reassuring the central role of dorsal striatum for the shared cognitive abnormalities of the psychopathology of schizophrenia and the clinical symptoms of schizophrenia.

Finally, we investigated univariate genetic variants shared between schizophrenia and subcortical Cog-ReFS. By associating subcortical ReFS (8624, 539*16) with 279 single nucleotide polymorphisms (SNPs) identified in a recent schizophrenia genome-wide association study (GWAS) (*26*), we found significant SNP-brain associations for 172 out of 279 SNPs (70%) after correction for multiple testing (**Fig. 3c**), with four top SNPs represented 46% of all significant SNP-brain associations (**Fig. 3c** and **Table S16**). In contrast, the subcortical volumes only have significant associations with 37 out of 279 SNPs (13%, **Table S17**). The associations of the four top SNPs showed different regional distributions (**Fig. 3d, Table S18)**. For instance, the T allele of rs13107325 had the greatest number of associations across all the 16 subcortical regions and was also one of the top risk genetic effects in the original schizophrenia GWAS (OR = 1.15, P = 2.99E-23) (*26*). The rs13107325 was associated with increased Cog-ReFS in pPUT for motor-related factors and “*illusion*” and also decreased Cog-ReFS in pPUT with “*feelings*”, “*language*”, and “*theory of mind*” **(Fig. 3e** and **Table S15)**. The above results hence confirmed highly overlapped genetic substrates between schizophrenia and subcortical brain regions using our ReFS approach.

## Discussion

The present study demonstrated the crucial role of dopamine-based striatal representational feature score (ReFS) in schizophrenic psychopathology, and remarkably, the increased striatal dopamine-ReFS in schizophrenic patients could be consistently reproduced across all 13 tested independent datasets. In contrast, previous large-sample structural studies rarely found consistent mean/variance-based volumetric alternations of striatum in chronic or antipsychotic-free schizophrenia patients (*27, 28*). Such a discrepancy could be explained by our observations that brain regions with higher dopamine density tend to have diminished case-control brain mean-based volume difference while, instead, showing increased differences in the corresponding dopamine-ReFS. Notably, this effect of increased striatal dopamine-ReFS was also evident in the antipsychotic-free schizophrenia subgroup and the first-degree healthy relatives of schizophrenia patients. Therefore, the increased striatal dopamine-ReFS were most unlikely the results of confounding effects from antipsychotic medications and could serve as endophenotypic biomarkers for the vulnerability of dopaminergic dysfunction in schizophrenia.

Further, with high-dimensional latent cognitive factors, we found that the structural Cog-ReFS abnormalities in schizophrenia were also primarily located in the striatum, particularly the dorsal part (i.e. the globus pallidus, GP and the putamen, PUT), which corroborates a recent proposition asserting the central role of the dorsal striatum in the pathophysiology of schizophrenia (*29*). The dorsal striatum has been proposed as an integrative hub that regulates information transactions between limbic and motor regions (*30*), most notably through the dopaminergic projections from SNc. Our results indicated that Cog-ReFS abnormalities of the dorsal striatum in schizophrenia could influence multiple latent cognitive domains (i.e., motor-related, decision-making, and social-emotional functions). Besides, in the predictive model for the three schizophrenia symptom subscales (i.e., positive, negative and general subscales from PANSS) using all subcortical Cog-ReFS, the dorsal striatum was the primary contributor. These findings supported that dorsal striatum dysfunction was not merely associated with motor deficits in schizophrenia but may also contribute to other psychopathologies (such as emotionlessness and delusion) in schizophrenia. Therefore, the dorsal striatal dopamine level could be a promising treatment target for schizophrenia (*9, 31*). In a recent clinical trial, a highly selective M4 positive allosteric modulator, i.e. responsible for dopamine release in the dorsal striatum, has demonstrated a favourable safety profile in schizophrenia treatment (*32*).

Previous studies have consistently found a surprisingly low genetic correlation between schizophrenia and brain subcortical mean-based volume (*22*), which, as suggested by a recent finding (*33*), is likely because subcortical mean-based volume and schizophrenia share plenty of common genetic variants but with heterogeneous effects directions. In the present study, ReFS of multiple subcortical regions demonstrated significant genetic correlations with schizophrenia, indicating that ReFS could help to capture a more homogenous genetic construct of brain structure shared with schizophrenia (*34*). In terms of common genetic variants, we demonstrated that the same SCZ risk alleles were simultaneously associated with increased positive-symptom Cog-ReFS (i.e. motor and hallucination) and decreased negative-symptom Cog-ReFS (i.e. emotion, social and language) (Figure 3E), and both groups of Cog-ReFS were also associated with schizophrenia diagnoses (Figure 2B). Hence, this latent representation-based brain pattern (i.e. the ReFS) could also provide valuable insights into the genetic construction of neural mechanisms underlying schizophrenia.

## Data Availability

All data produced in the present study are available upon reasonable request to the authors

## Acknowledgements

None

## Author contributions

Conceptualization: C.X., T.J.T., F.F.J.; Methodology: C.X., T.J.T.; Formal analysis: C.X.; Writing: original draft: C.X., T.J.T.; Writing-review and editing: C.X., T.J.T, O.H., G.S., T.R.

## SUPPLEMENTARY MATERIALS

**Materials and Methods Figs. S1 to S14**

**Tables S1 to S20**

## ACKNOWLEDGMENTS

COBRE (The Center for Biomedical Research Excellence in Brain Function and Mental Illness) has published structural and functional MRI data obtained from schizophrenia patients and normal subjects. MCICShare comprised structural and resting-state functional MRI data, which was collected and shared by the Mind Research Network and the University of New Mexico funded by a National Institute of Health Center of Biomedical Research Excellence (COBRE) grant 1P20RR021938-01A2. Research using Human Connectome Project for Early Psychosis (HCP-EP) data reported in this publication was supported by the National Institute of Mental Health of the National Institutes of Health under Award Number U01MH109977. The HCP-EP 1.1 Release data used in this report came from DOI: 10.15154/1522899. This work received support from the following sources: the National Natural Science Foundation of China (T2122005 [to TJ], 82150710554 [to GS], KRH2306051 [CX] and 81801773 [to TJ]), China Postdoctoral Science Foundation (no. BX20230075 and 2023M740659 to CX). MOE Frontiers Center for Brain Science [to CX], National Key R and D Program of China (2023ZY1068 [to TJ], 2019YFA0709501 [to TJ], 2021YFC2501402 [to TJ], 2019YFA0709502 [to JF], 2018YFC1312900 [to TJ] and 2018YFC1312904 [to JF]), the Shanghai Pujiang Project (18PJ1400900 [to TJ]), Guangdong Key Research and Development Project (No. 2018B030335001 [to JF]), the European Union-funded FP6 Integrated Project IMAGEN (Reinforcement-related behavior in normal brain function and psychopathology) (LSHM-CT-2007-037286 [to GS]), the Horizon 2020-funded ERC Advanced Grant’ STRATIFY’ (Brain network based stratification of reinforcement-related disorders) (695313 [to GS]), the 111 Project (B18015 [to JF]), the key project of Shanghai Science and Technology (16JC1420402 [to JF]), Shanghai Municipal Science and Technology Major Project (no. 2018SHZDZX01 [to JF]), ZJ Lab [to JF], Shanghai Center for Brain Science and Brain-Inspired Technology [to JF], ERANID (Understanding the Interplay between Cultural, Biological and Subjective Factors in Drug Use Pathways) (PR-ST-0416-10004, [to GS]), Human Brain Project (HBP SGA 2, 785907, and HBP SGA 3, 945539, [to GS]), the Medical Research Council Grant’ c-VEDA’ (Consortium on Vulnerability to Externalising Disorders and Addictions) (MR/N000390/1 [to GS]), the National Institute of Health (NIH) (R01DA049238, A decentralised macro and micro gene-by-environment interaction analysis of substance use behavior and its brain biomarkers [to GS]), the National Institute for Health Research (NIHR) Biomedical Research Centre at South London and Maudsley NHS Foundation Trust and King’s College London, the Bundesministeriumfur Bildung und Forschung (BMBF grants 01GS08152; 01EV0711 [to GS]; the European Union and UKRI funded project environMENTAL (grants 101057429 [to GS]). Forschungsnetz AERIAL 01EE1406A, 01EE1406B [to GS]), Forschungsnetz IMAC-Mind 01GL1745B [to GS]), the Deutsche Forschungsgemeinschaft (DFG grants SM 80/7-2, SFB 940, TRR 265, NE 1383/14-1, [to GS]).

