## supplementary figure for "Dopaminergic-related Anatomical Pattern of Dorsal Striatum in Schizophrenia"

**The PDF file includes:**

Materials and Methods

Fig S1-S12

Reference

**Materials and Methods**

Datasets & Subjects

Six datasets were used in this study, including the SIMON dataset(*1*), Myconnectome dataset (*2*), Donders-PET (*3*), SRPBS (*4*), Schizophrenia clinical datasets(*5*), and UKB dataset(*6*).

**SIMON** **dataset.** The Single Individual volunteer for Multiple Observations across Networks (SIMON) MRI dataset is a sample of one healthy male aged between 29 and 46 years old, scanned with 49 times at multiple sites and with various scanner models.

**Myconnectome dataset.** MyConnectome data contains 21 times of magnetic resonance imaging scans of a 45 years old healthy male recorded within an 18 month period.

**Donders-PET**. This collection shares raw data from a project comprising multiple subprojects targeting the development of a dopamine proxy-model using machine learning (namely Donders-PET dataset). In a double-blind, placebo-controlled design, 100 Dutch speakers underwent sessions involving pharmacological interventions, fMRI, and [18F]-fluoro-dopa injections for PET scans.

**SRPBS.** In the Japanese Strategic Research Program for the Promotion of Brain Science (SRPBS) travelling subject dataset, 9 participants (all male; age range 24–32y; mean age 27±2.6 y) individually scanned in 12 different scanners, totally 108 sMRI scans, which was used for sensitivity analysis (Fig S).

**Schizophrenia clinical datasets.** The study contains 1,229 schizophrenia patients and 1,237 healthy controls form 13 datasets, including ZhangJiang International Brain Biobank (ZIB, dataset #1, dataset #2), Taipei Veteran General Hospital (Taipei, dataset #3), First Affiliated Hospital of Zhengzhou University (dataset #4), Xijing Hospital (Xijing, dataset #5), Clinical Hospital of Chengdu Brain Science Institute in Chengdu (ChengDu, dataset #6) and from another 7 publicly available datasets, i.e., JapanKUT (dataset #7), JapanUTO (dataset #8), JapanKTT(dataset #9), DS000115 from OpenfMRI database (WM, dataset #10), Human Connectome Project for Early Psychosis dataset (HCP-EP, dataset # 11), The Mental Illness and Neuroscience Discovery Institute (MIND) Clinical Imaging Consortium (MCIC, dataset #12), The Center for Biomedical Research Excellence dataset (COBRE, dataset #13), NMorphCH (dataset #6), FBIRN (dataset #7), NUSDAST (dataset #8) and DS000115 (dataset #9). All individuals with schizophrenia were diagnosed according to the Diagnostic and Statistical Manual of Mental Disorders, 4th Edition (DSM-IV). The study protocol was approved by the Medical Research Ethics Committees of the local hospitals and written informed consent was obtained from all participants and/or their legal guardians. Individuals with illness duration less than 2 years were defined as first-episode schizophrenia (FES)(*7*).

**UKB**. We used data from the UK Biobank (UKB) with application ID 19542. The UK Biobank has research tissue bank approval from the North West Multi-centre Research Ethics Committee (https://www.ukbiobank.ac.uk/learn-more-about-uk-biobank/about-us/ethics) and provided oversight for this study. Written informed consent was obtained from all participants. Participation is voluntary, and participants are free to withdraw at any time without giving any reason. Neuroimaging data of 34,497 participants were available and used in the current analyses under the application number 19542. All participants provided written informed consent. Quality-controlled T1-weighted neuroimaging data were used in current study. Details of the imaging protocol can be found in an open-source document (https://biobank.ndph.ox.ac.uk/showcase/showcase/docs/brain_mri.pdf). Neuroimaging data were collected with a standard Siemens Skyra 3T scanner with a 32-channel head coil.

Third-party functional neuroimaging datasets

Two public available group-level functional neuroimaging datasets were used in present study: **a neurotransmitter maps dataset** from the tool of JuSpace with nine neurotransmitter maps, involving various neurotransmitter systems, such as dopaminergic, serotonergic, noradrenergic, and GABAergic (gamma-aminobutric acid) neurotransmission (*8*); **a task-based meta-analysis maps dataset** from the website of *Neurosynth* (*9*), which have plenty of cognitive-behavior meta analytic activation maps from primary information processing to high-order cognitive functions. According to a previous study(*10*), we selected 539 cognitive-behavior maps, such as reward processing, social emotion, and decision making. The two types of group-level maps were used as function reference to estimate individual-level brain spatial pattern similarity for each subject.

T1-weighted MRI data preprocessing

T1-weighted MRI acquisition parameters of these datasets have been provided previously (*1, 2, 4-6*). We utilized the Computational Anatomy Toolbox (http://www.neuro.uni-jena.de/cat/) within SPM12 (<https://www.fil.ion.ucl.ac.uk/spm/software/spm12/>) to processed these T1-weighted images. Specifically, the standard voxel-based morphometry (VBM) (including spatial registration, tissue segmentation and bias correction of intensity non-uniformities, smoothed at 8 mm FWHM) was conducted, generating voxel-level gray matter volume (GMV) images. Then, a recently published subcortical template (*11*) was used to extract each regional voxel-level GMV values for each subject and then used for following analyses.

Structural representational feature analysis with functional reference

We proposed a novel computational framework to decompose regional spatial pattern of grey matter volume with the priori biological information. Specifically, we firstly normalize the grey matter volume of each brain region to avoid the inter-individual heterogeneity effect (*12*); Then, we calculated the covariance between the regional grey matter volume and functional reference maps for each participant. Compared to the univariate-based measurements of mean and variance, this weighted approach helps to extracted more specific biological information from structural imaging, generating individual-level representation feature score (ReFS) for each brain region with different functional reference.

Reliability analysis of brain ReFS

We also investigated the reliability of third-party reference with the spatial map of F-DOPA as an example. Utilizing the Donders-PET dataset, we kept 94 participant data for following analyses after quality control. We used two approaches to evaluate the reliability of the group-level F-DOPA spatial maps (**Supplementary Fig. 3**). First，we checked the spatial correspondence between the two group-level F-DOPA spatial maps that which from the Donders dataset and JuSpace. The P value of spatial similarity was estimated with a standard non-parametric null model (*13*). We found that the two group-level F-DOPA maps from different datasets showed high significant correlation (*r* = 0.94, *P_shuf_* = 0). Next, we used the bootstrap sampling to generate 1000 null group-level F-DOPA maps with the Donders dataset. The spatial similarity between the null group-level F-DOPA maps and JuSpace F-DOPA map was range from 0.94 and 0.95. The results suggested that the spatial pattern of neurotransmitter is highly consistent across sites and groups.

Reproducibility analysis of brain ReFS

We estimated the test-retest reproducibility of brain ReFS with intraclass correlation coefficient (ICC), which was commonly used in neuroimaging study (*14*). Statistically, the reproducibility of measure provide an upper bound on the possible correlation that can be observed with the measure (*15*). The ICC values range from 0 to 1, with higher values indicating better reproducibility. A value of 0 indicates no agreement among repeated measurements, while a value of 1 indicates perfect agreement. There are three grade of ICC value that ICC lower 0.4 means poor reproducibility, ICC between 0.4 and 0.75 means fair to good reproducibility, and ICC larger 0.75 means excellent reproducibility. As a benchmark in neuroimaging(*16*), we also estimated the ICC values of the brain volume to check whether brain ReFS help to improve the reproducibility of the measurements of structural imaging. The two repeated density scan datasets of SIMON and Myconnectome were used in the present study. This detailed identification procedure was illustrated in **Supplementary Fig. 4 and 5.**

Sensitivity analysis of brain ReFS

We used the brain fingerprint analysis to check whether the brain ReFS was sensitive to detect the individual differences with the SRPBS travelling participants dataset, in which nine participants were scanned in 12 different scanners. The brain fingerprint analysis was first introduced to show that functional connectivity profiles act as a ‘fingerprint’ that can accurately identify subjects from a large group. Specifically, first, a target database was constructed that including all the participants’ informative pattern matrices from one scanner. Next, for the subject identification, the similarity was computed with Pearson correlation between the target database and one subject brain ReFS matric from another scanner. This identification process was conducted interactively for the 12 scanners. The fingerprint analysis was also conducted for brain mean-based volume. This detailed identification procedure was illustrated in **Supplementary Fig. 6.**

Group comparison between schizophrenia patients and healthy controls

In the present study, we collected 1,229 schizophrenia patients (SCZ) and 1,237 healthy controls (HC) from 13 datasets. The liner mixed models (LMM) was utilized to investigate group difference of subcortical ReFS and mean-based volume/variance between SCZ and HC, while allowing site-varying effects. LMM estimates the relationship between a response variable (e.g., ReFS of F-DOPA-Putamen) and independent variables (here, diagnosis and covariates of age, sex, and total intracranial volume (TIV)), with coefficients that can vary with respect to grouping variables (here, site) (*17, 18*). We used MATLAB’s command fitlme (https://www.mathworks.com/help/stats/fitlme.html) to estimate the model: y ∼1 + Diagnosis + Age + Sex +(1 | Site) + (Diagnosis | Site), which yields *t* and *P* values for the fixed effect of Diagnosis.

The association analysis between group difference and neurotransmitter level at region-level

With the LMM model, we estimated the group difference for each subcortical region for brain ReFS and mean-based volume. We then asked whether the variance of group difference between subcortical regions was modulated by the neurotransmitter level. We compared the regional group difference to regional neurotransmitter levels using the Kendall rank correlation coefficient (Kendall τ).

The association analysis between Cog-ReFS group difference and neurotransmitter relevance.

As neurotransmitter underlie the cognitive functions, we asked whether Cog-ReFS group difference between SCZ and HC was modulated by neurotransmitter by investigating the Pearson correlation between regional-level Cog-ReFS group difference and the neurotransmitter relevance between cognitive maps and neurotransmitter maps (**Supplementary Fig. 9 a, b and c**). The significance was estimated with permutation test (**Supplementary Fig. 9 d**). Specifically, in each permutation iteration, we reconducted the previously mentioned group comparison using randomly reshuffled participant labels and estimated the correlation between Cog-ReFS group difference and neurotransmitter relevance.

The predictive analysis between structural Cog-ReFS and schizophrenic symptom severity

With the 539 cognitive-behavior maps, the regional grey matter volumes were mapped into high-dimensional space that each subcortical region would have 539 different ReFS, such as motor response, emotional preprocessing, and language etc. For the subcortex, we generated a high-dimensional Cog-ReFS matrix (16*539, 16 regions, 539 cognitive terms). We hypothesized that this high-dimensional structural ReFS offered a unique chance to predict the schizophrenic symptom severity.

We employed connectome-based predictive modeling (CPM) (*19*) to predict the participants' schizophrenia symptoms from high-dimensional subcortical Cog-ReFS. CPM is a recently developed method for identifying functional brain connections related to a behaviour variable of interest, such as fluid intelligence, attention control and ADHD (*20-23*). The CPM pipeline is available online (<https://www.nitrc.org/projects/bioimagesuite/>).

Here, we extended this method to explore whether high-dimensional brain Cog-ReFS could predictive the schizophrenia symptoms with the leave-one-out cross-validation. Specifically, In the first step, we select one subject as the test dataset, and the other (N-1) subjects as the training dataset. Next, a vector of behavioural scores (e.g. schizophrenia symptoms) was associated with the brain Cog-ReFS (i.e. the Cog-ReFS matrix, 16*539) across participants from the training dataset, with site and handedness, gender being included as covariates. Then, a default threshold (*19*) (i.e. P < 0.01 in the present study) was applied to retain only ReFS that were significantly associated (either positively or negatively) with behavioural symptoms in the training dataset. Next, the sum of positive and negative ReFS’ weights (negative edges will be multiplied by -1 before sum-up) was calculated for each individual and then entered into a linear regression model to estimate the relationship between the summed ReFS score and the observed behaviour in the training dataset. In the testing dataset, the summed ReFS score of each individual was submitted to the corresponding linear model estimated in the training dataset to generate the predicted behaviour score. The above process was repeated for each subject, with predicted behaviour scores in each testing fold established based on the rest subject data. Finally, Spearman’s correlation was applied to estimate the model performance between predicted and actual behaviour scores across all individuals. Only brain ReFS selected in over 95% of models to select as the robust predictive feature for the following analyses.

The association analysis between Cog-ReFS group difference and Cog-ReFS predictive weights.

The high-dimensional brain ReFS showed significant group difference and predictive effect for symptom severity. We further asked whether they shared an similar variational spectrum across brain Cog-ReFS. We investigated the Pearson correlation between regional-level Cog-ReFS group difference and predictive weights for each subcortical region (**Supplementary Fig. 11 a, b and c**). The significance was estimated with permutation test (**Supplementary Fig. 11 d**). Specifically, in each permutation iteration, we reconducted the previously mentioned group comparison using randomly reshuffled participant labels and estimated the correlation between Cog-ReFS group difference and predictive weight for each subcortical region.

Genotyping and quality controls

The imputed genetic variants data was download from UKB data resource. We next conducted following genetic variants data quality controls: i) removed subjects with more than 10% missing genotypes; ii) removed variants with minor allele frequency less than 0.01; iii) removed variants with missing genotype rate larger than 10%; iv) removed variants that failed the Hardy-Weinberg test at 1 × 10^−7^ level.

Heritability analysis of brain representational features

We estimated the single-nucleotide polymorphism (SNP)-based heritability (*h*^2^) for the informative pattern of subcortical regions with the UKB individuals of white British ancestry (N = 34,496) by genome-wide complex trait analysis (GCTA) (*24*). We controlled the following confounding variables: age (at imaging), age-squared, sex, age–sex interaction, age-squared–sex interaction, imaging site, head location, head motion, head size, long-term drifts and the top 40 genetic PCs. We used the ‘—grm-cutoff 0.025’ option in GCTA to remove individuals with estimated relatedness larger than 0.025.

Genome-Wide Association Studies (GWAS)

Genome-wide association analysis was conducted with fastGWA(*25*), which was commonly used in the imaging-genetics studies for high-dimensional neuroimaging data (*26*). The main GWAS of brain ReFS patterns was performed on the British individuals (self-reported ethnic background, Data-Field 21000) in UK Biobank study. The same confounding variables used in the heritability analysis were also controlled in the brain ReFS GWAS. Related individuals were included in fastGWA, and linear mixed-effect model-based approaches were used to account for the sample relatedness.

Genetic correlation

We applied both the LD score regression(*27*) and high-definition likelihood methods(*28*) to evaluate the genetic correlations between subcortical imaging phenotypes and schizophrenia. GWAS summary statistics were harmonized by the munge_sumstats.py procedure in the ldsc software. The intercept score in the LDSC analysis were used to check whether there is genomic inflation effect caused by confounding factors.

**Supplementary Figures**

**Fig S1. Demonstration of the ReFS**

**Fig S2. Overview of research question and analyses**

**Fig S3. The reliability analysis of ReFS with the Donders-PET dataset**

**Fig S4. The reproducebility analysis of ReFS with the Myconnectome dataset**

**Fig S5. The reproducebility analysis of ReFS with the SIMON dataset**

**Fig S6 The sensitivity analysis of ReFS with the SRPBS dataset**

**Fig S7. The spatial distribution of neurotransmitter maps**

**Fig S8. The spatial distribution of task-based meta-analysis activation maps**

**Fig S9. The association between group difference and neurotransmitter relevance**

**Fig S10. The predictive analysis with modified CPM**

**Fig S11. The association between predictive weights and neurotransmitter relevance**

**Fig S12. The heritability analysis of brain ReFS**

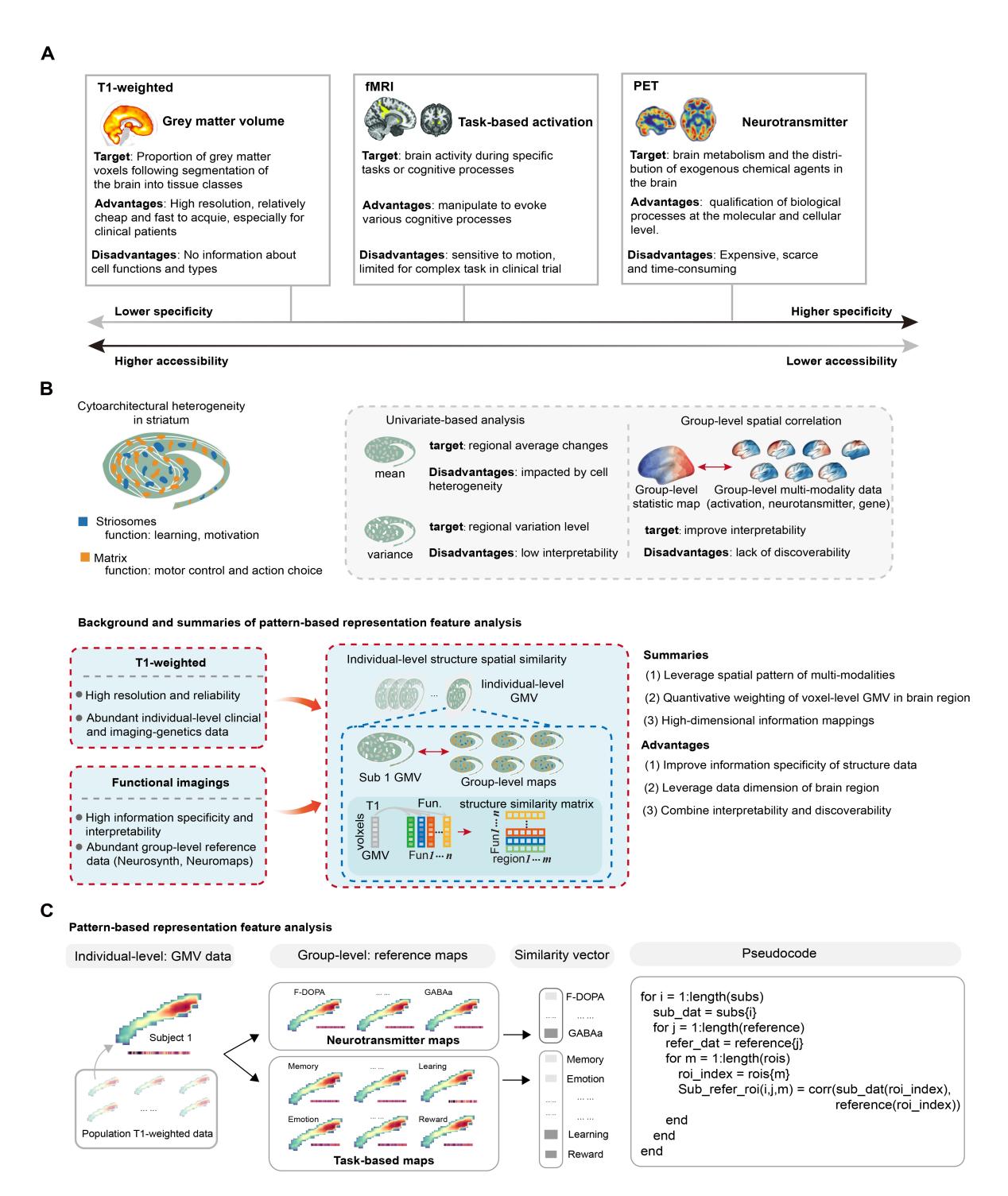

Fig S1. Demonstration of the ReFS. **a**. the specificity line is not linear but illustrates the rank order of specificity for neuron. **c**. the illustration of pattern-based the representation feature analysis with third-party group-level functional maps.

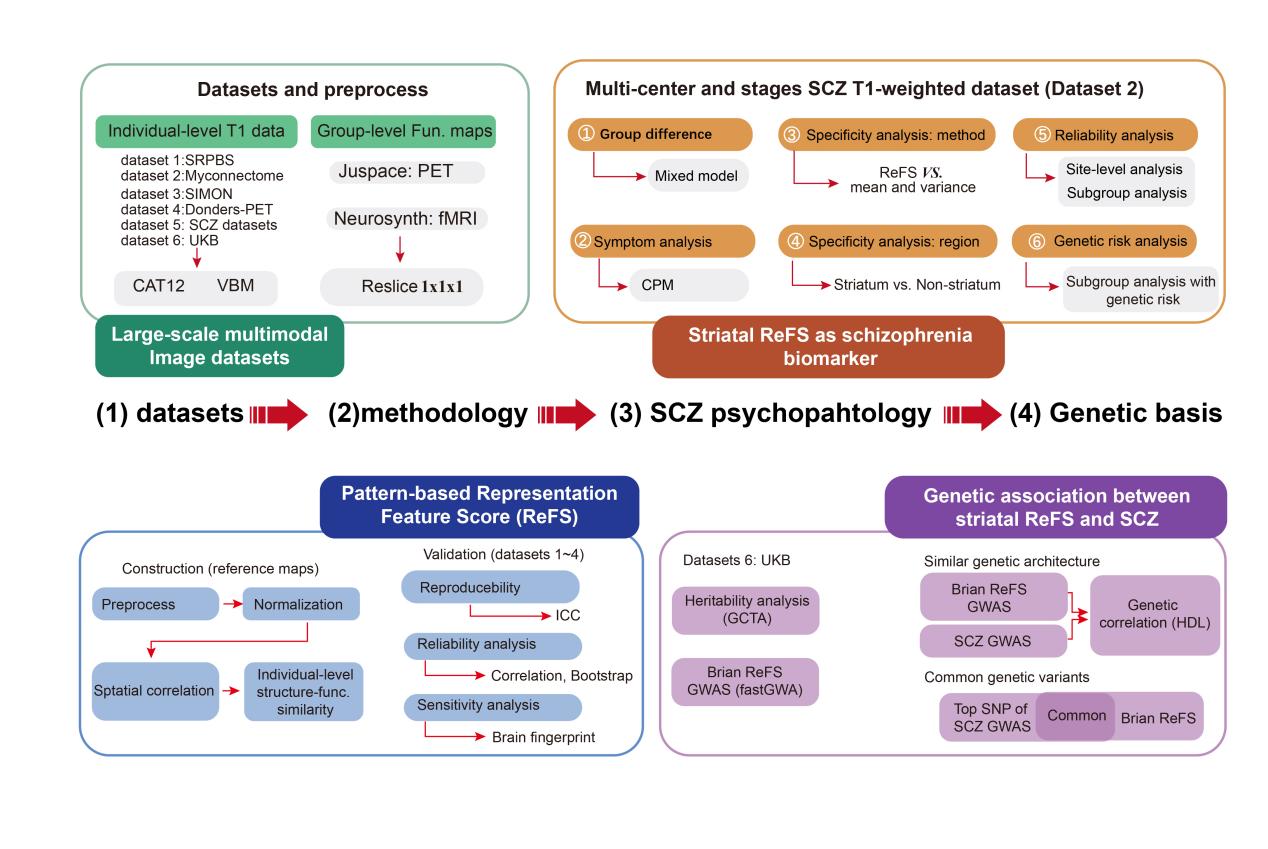

Fig S2. Overview of research question and analyses

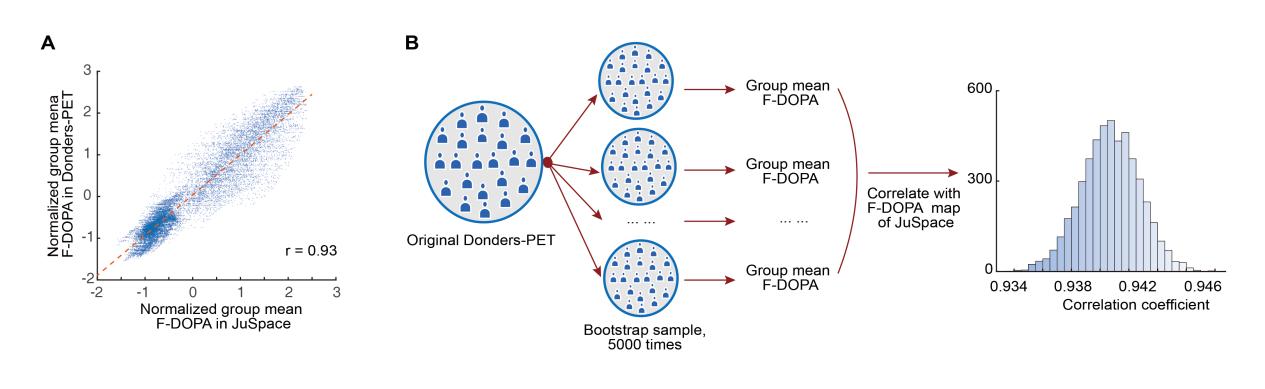

Fig S3. The reliability analysis of ReFS with the Donders-PET dataset. **a**. The spatial correlation between group-level F-DOPA maps of the JuSpace and Donders-PET datasets. **b.** Using the bootstrap 5000 times, we estimated the spatial correlation distribution between the 5000 group-level F-DOPA maps with the Donders-PET dataset and the group-level F-DOPA maps of the JuSpace.

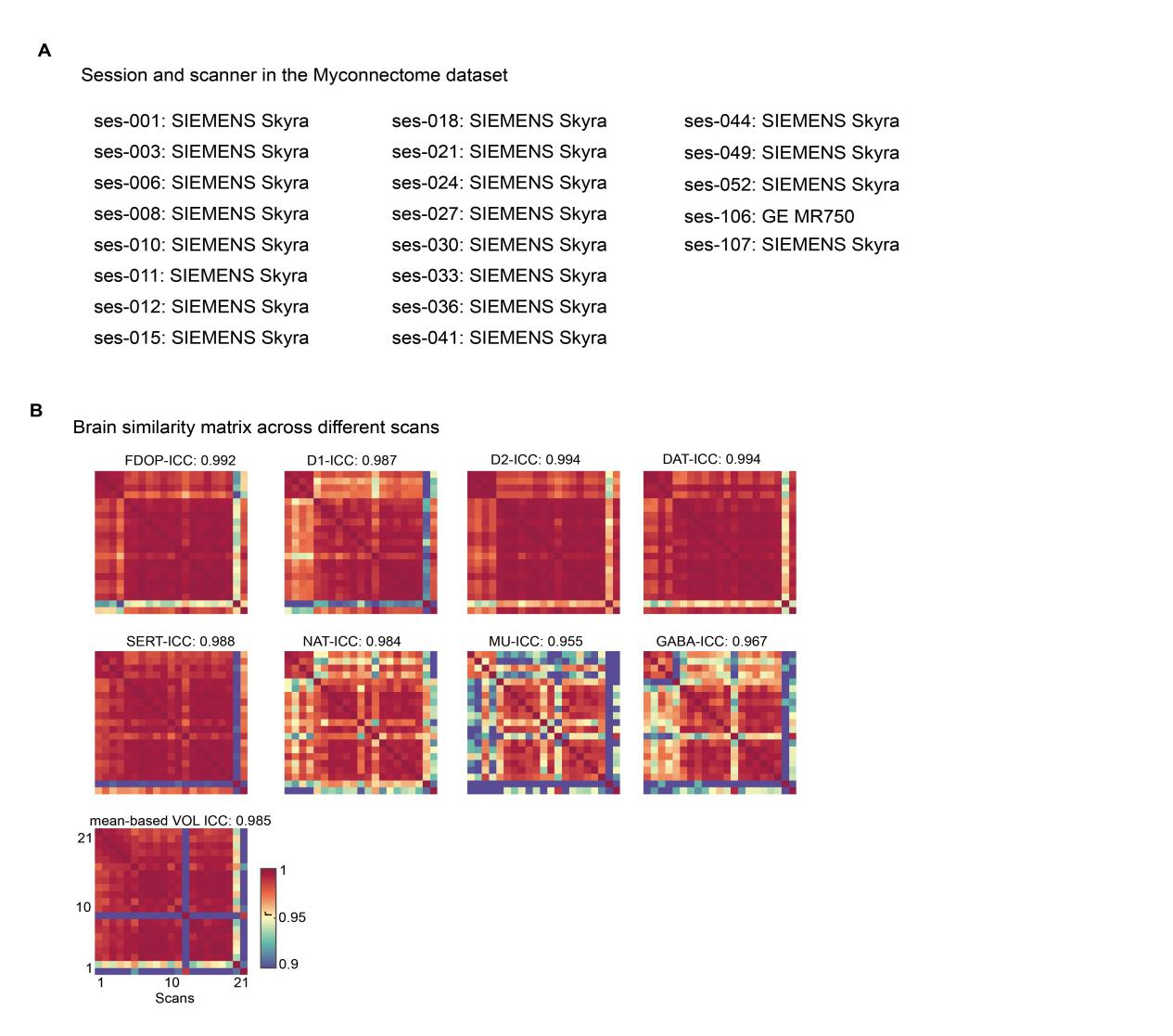

Fig S4. The reproducebility analysis of ReFS with the Myconnectome dataset. **a.** The details of data sessions and scanners in the Myconnectome datasets. After quality control, we only used 21 sessions in the present study. **b**. The intraclass correlation coefficient (ICC) and brain similarities between different scans with mean-based volume and PET-ReFS.

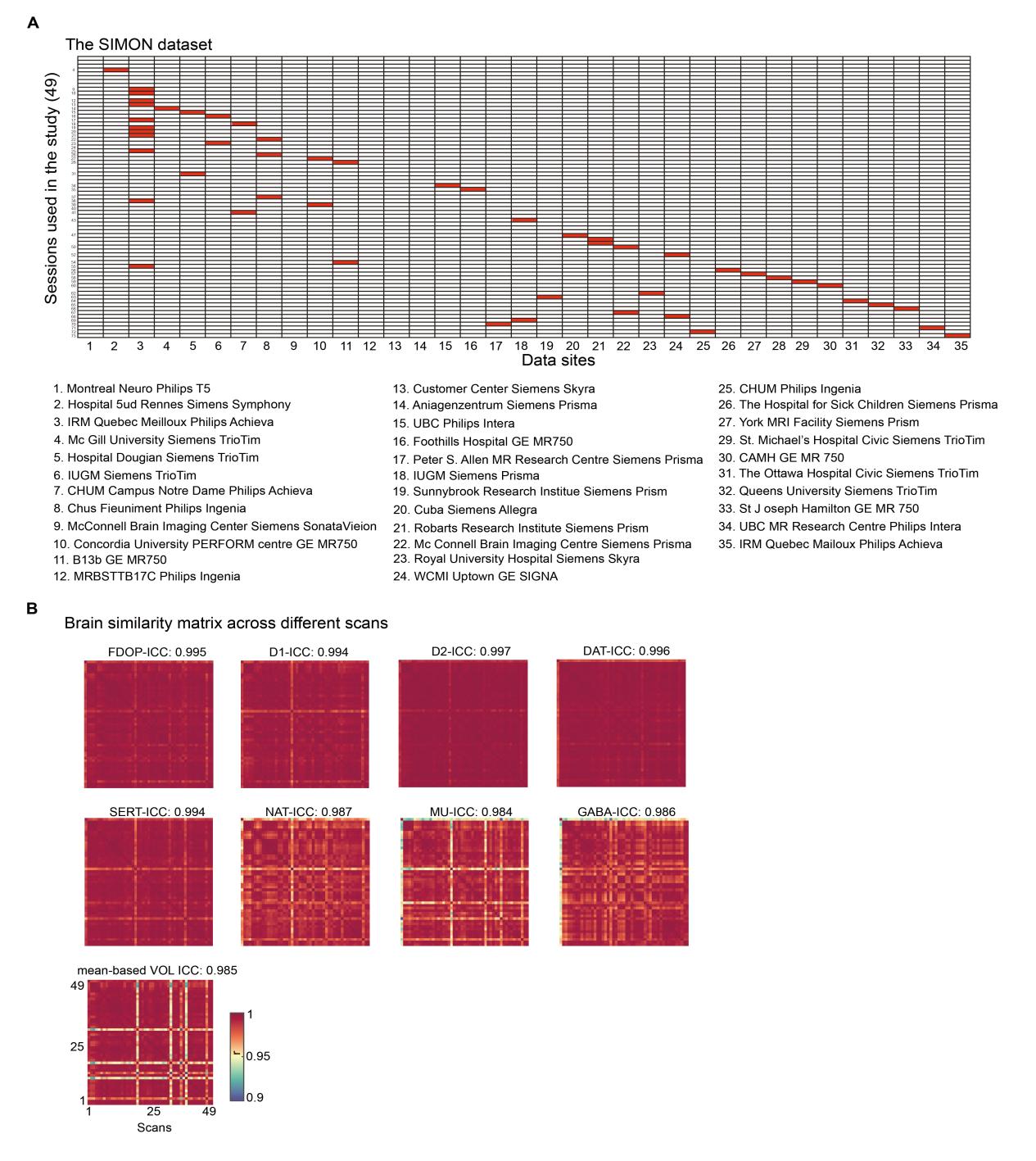

Fig S5. The reproducebility analysis of PET-ReFS with the SIMON dataset. **a**. The details of data sessions and scanners in the SIMON datasets. After quality control, we only used 49 sessions in the present study. **b**. The intraclass correlation coefficient (ICC) and brain similarities between different scans with mean-based volume and PET-ReFS.

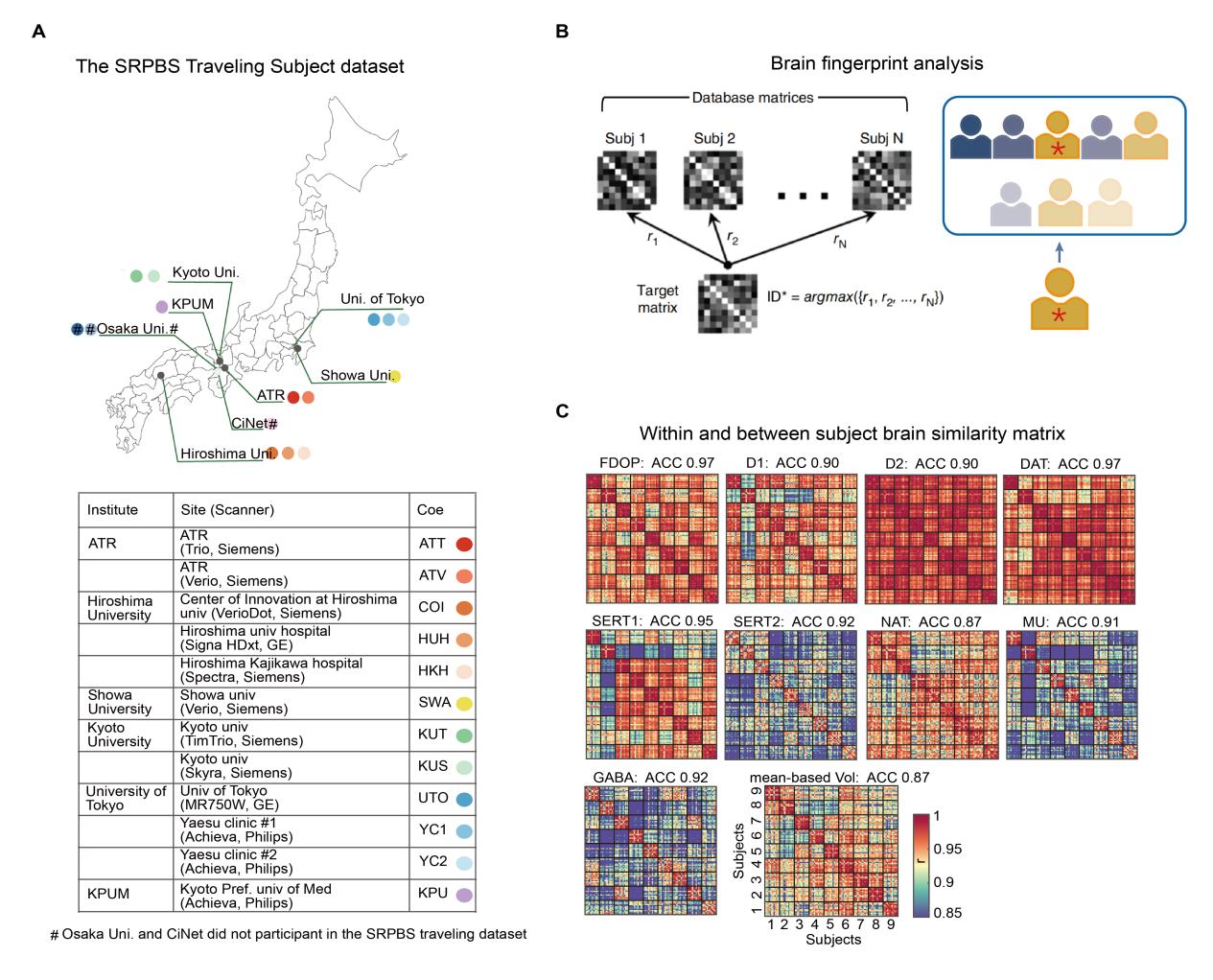

**Fig S6** The sensitivity analysis of ReFS with the SRPBS dataset. **a**. The sites, scanner and institute information of the SRPBS travelling dataset. **b.** the graphical illustration of the brain fingerprint analysis. **c**. The results of the brain fingerprint analysis and subject-level similarity matrix with mean-based volume and PET-ReFS. ACC, accuracy; Vol, volume. SRPBS, the Japanese Strategic Research Program for the Promotion of Brain Science.

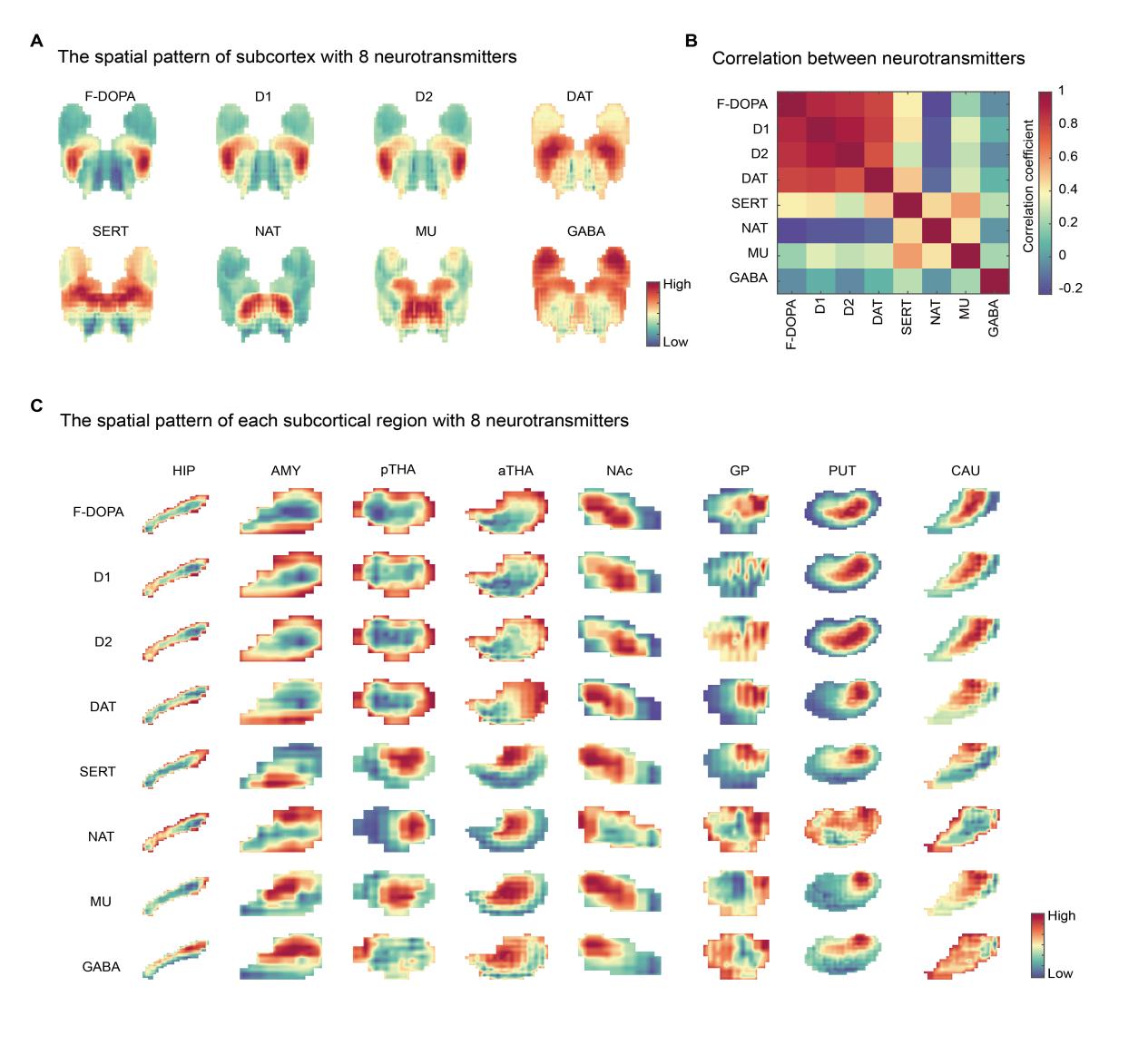

**Fig S7.** The spatial distribution of neurotransmitter maps. **a**. The spatial pattern of subcortex with 8 neurotransmitter maps. **b.** The spatial correlation between 8 neurotransmitter maps. **c.** The spatial pattern of each subcortical brain region with 8 neurotransmitter maps.

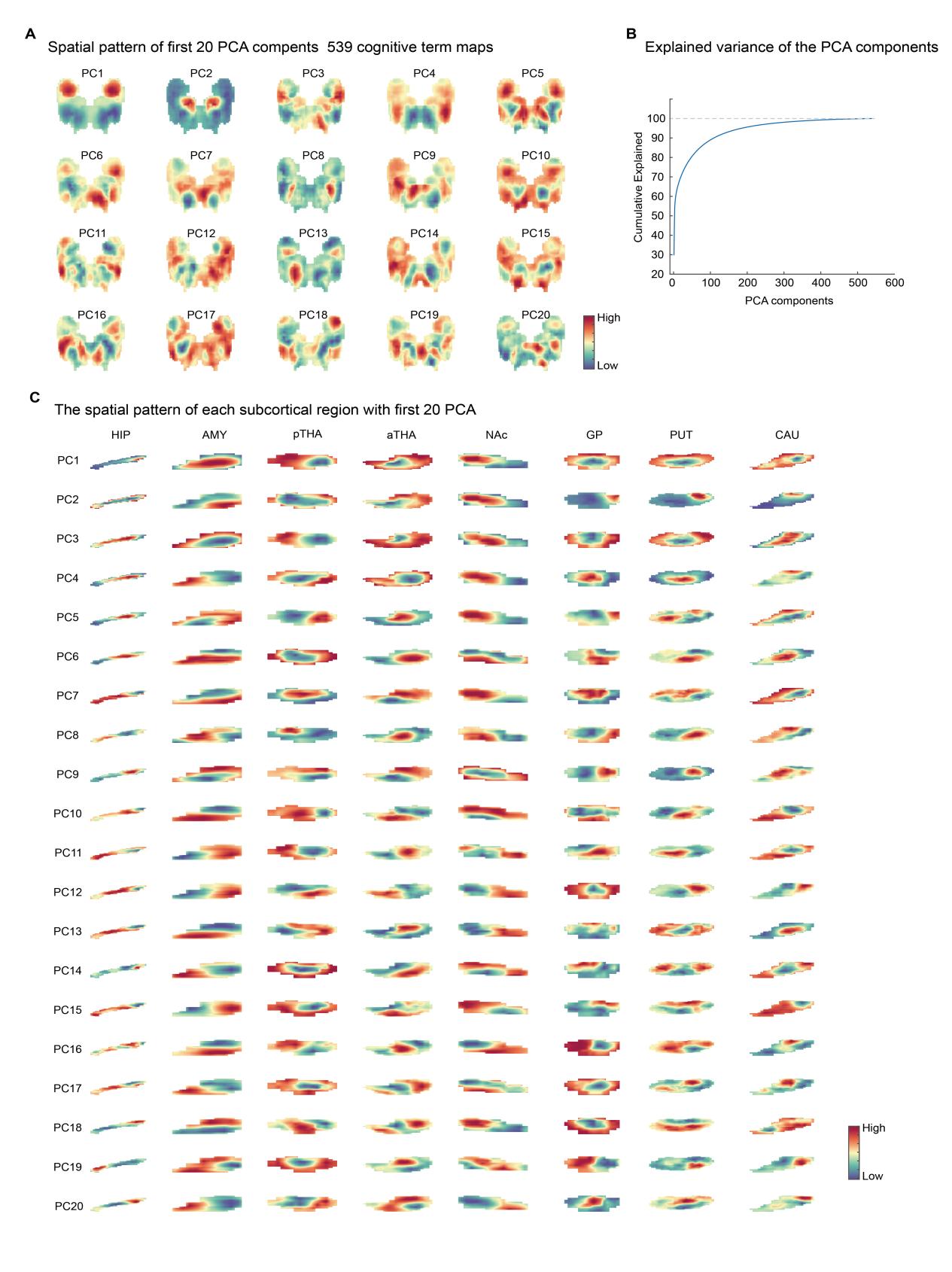

**Fig S8**. The spatial distribution of task-based meta-analysis activation maps. **a**. The spatial pattern of first 20 principal components (PC) with 539 cognitive maps. **b.** The cumulative explained variance line of principal components. **c.** The spatial pattern of each subcortical brain region with first 20 principal components.

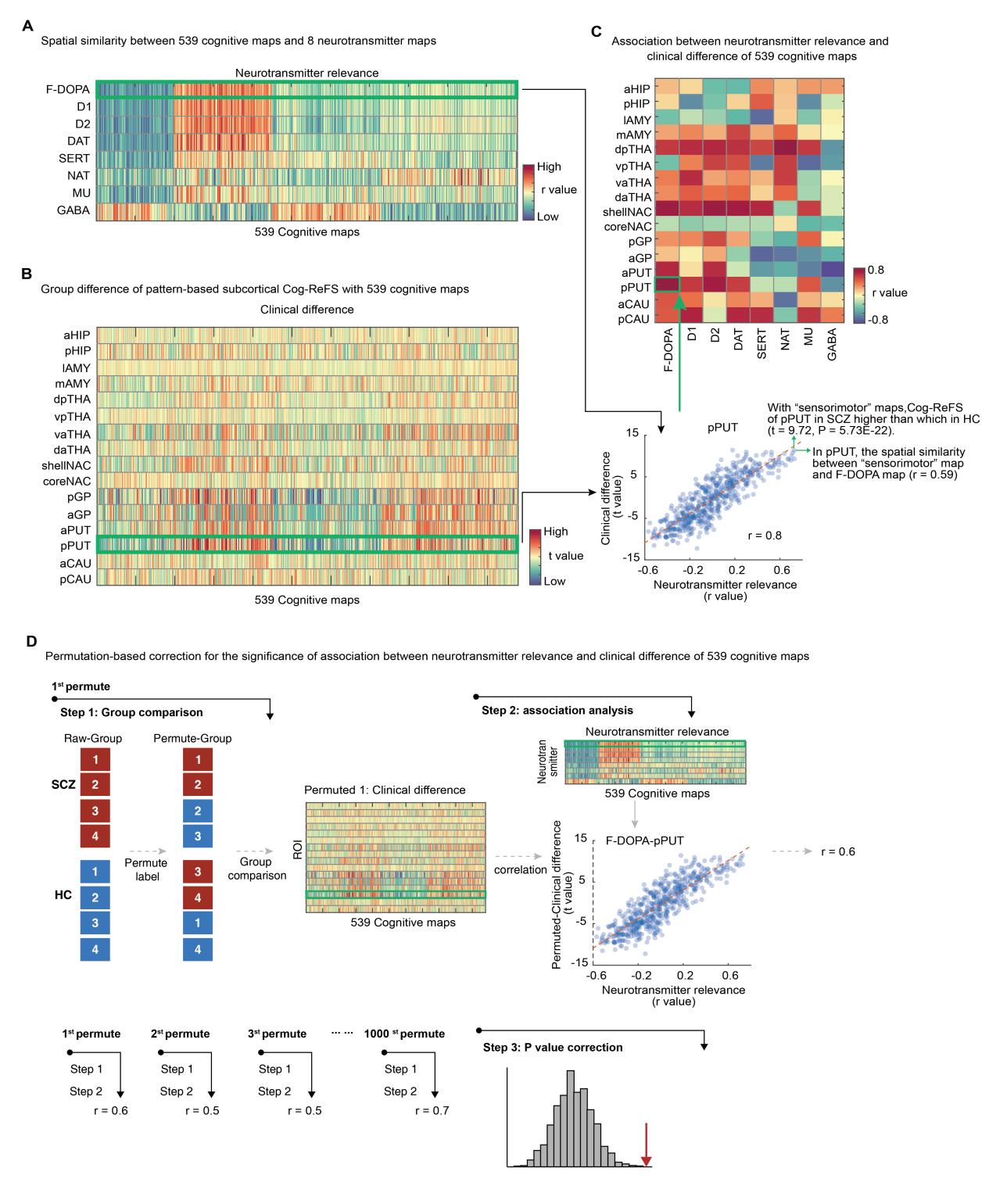

**Fig S9**. The association between group difference and neurotransmitter relevance

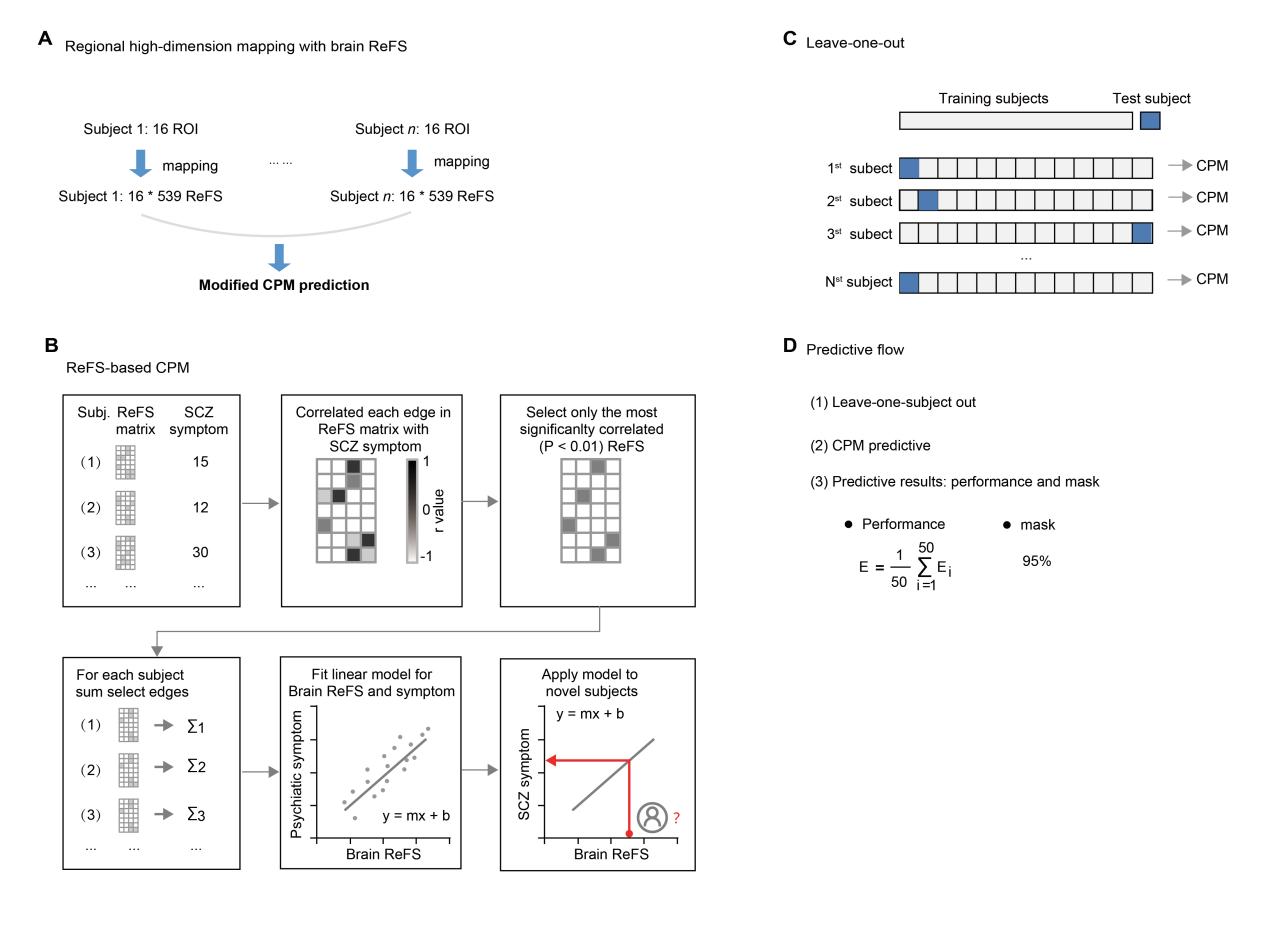

**Fig S10**. The predictive analysis with modified CPM. **a.** With high-dimensional brain-ReFS, we used the modified connectome-based predictive model (CPM) to predict the schizophrenia symptoms. **b.** The overview steps of the modified connectome-based predictive model. **c.** Leave-one-out cross validation. **d.** The predictive flow of ReFS-based CPM.

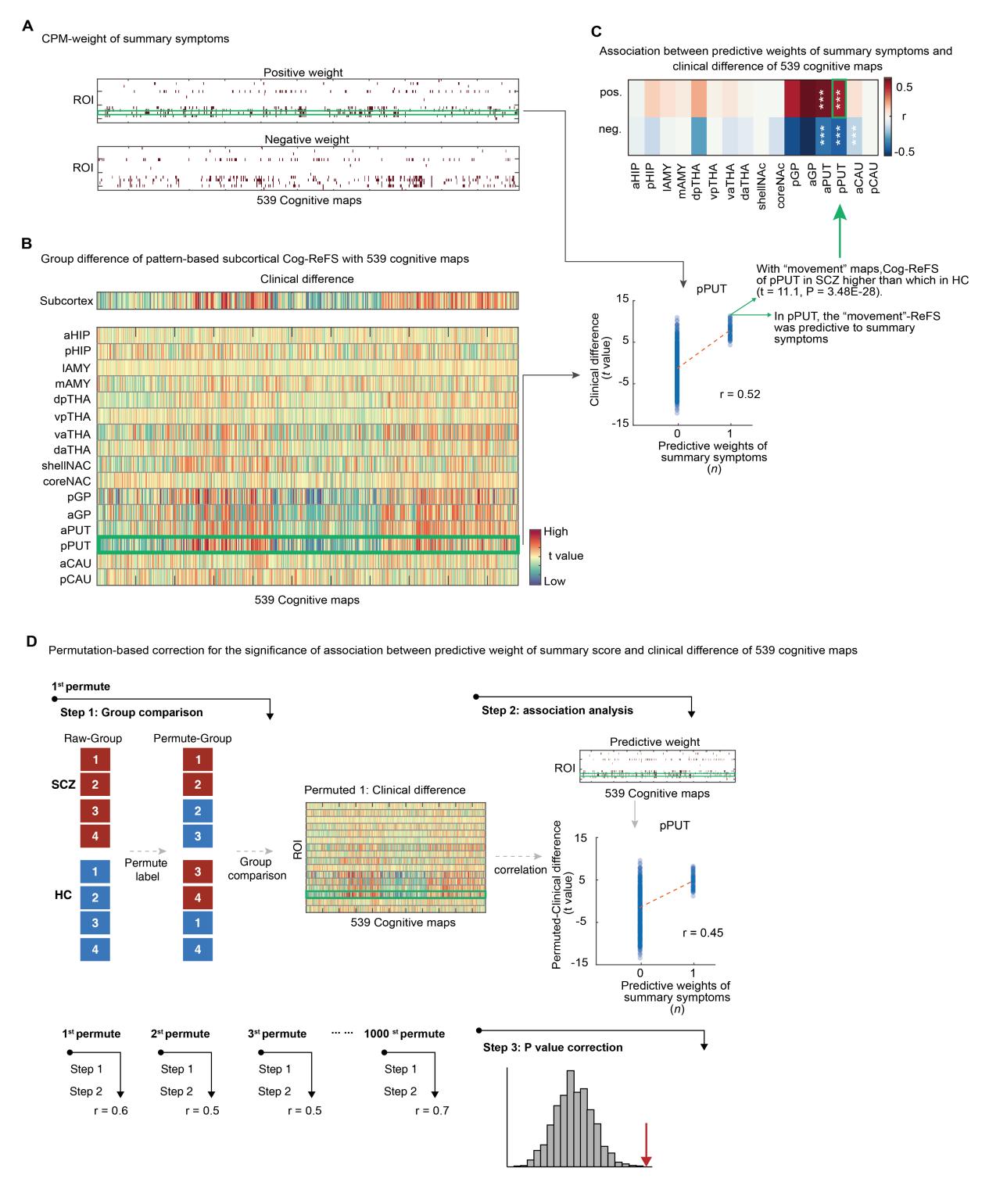

Fig S11. The association between predictive weights and neurotransmitter relevance

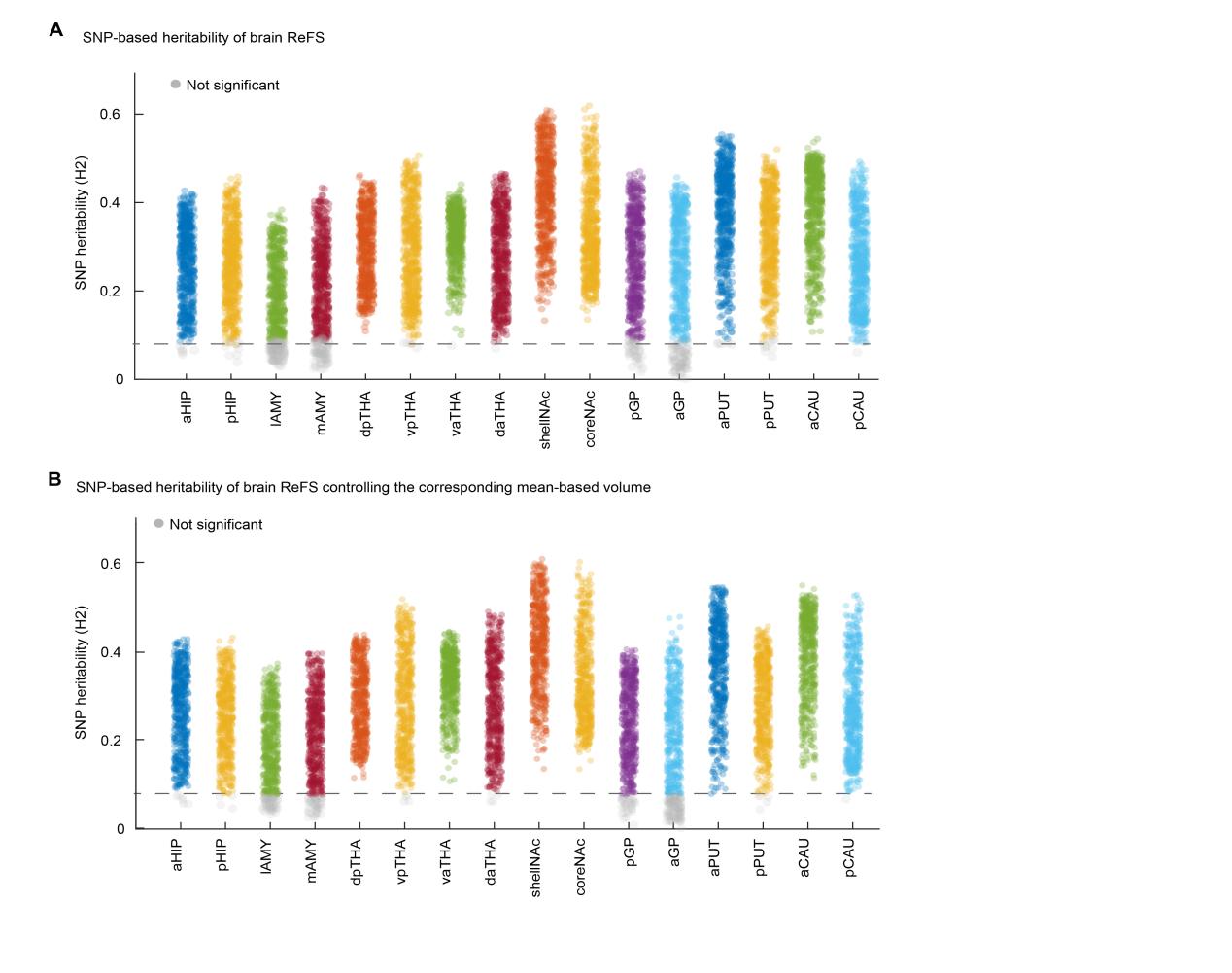

Fig S12. The heritability analysis of brain ReFS.
